# Big Five Personality Traits Influence Tinnitus Improvement Over Time

**DOI:** 10.1101/19000646

**Authors:** Jorge Simões, Winfried Schlee, Martin Schecklmann, Berthold Langguth, Daria Farahmand, Patrick Neff

## Abstract

Previous studies investigating the relation between personality and tinnitus distress showed that high neuroticism and low extraversion scores are related to higher tinnitus distress measured by the Tinnitus Handicap Inventory (THI) and the Tinnitus Questionnaire (TQ). However, little is known about the role of personality on tinnitus distress over time.

We collected the THI, TQ and Big Five Factor Index 2 of 388 patients who visited the Tinnitus Center Regensburg between 2012 and 2017, and filled in a survey with the same questionnaires in 2018. We used personality traits and facets to predict tinnitus distress cross-sectionally and longitudinally. Neuroticism and extraversion were significant predictors of THI and TQ scores in cross-sectional linear regression setups, and could explain up to 40% of the variance. However, the linear regressions could explain only little variance of both THI and TQ longitudinally. We clustered patients in three groups based in the difference THI and TQ between the two assessments: “clinically improved”, “clinically stable” and “clinically worsened”. The patients in the “clinically improved” and “clinically stable” groups scored statistically significantly lower in neuroticism and higher in extraversion than patients in the group “clinically worsened”. We observed a similar trend among patients who tried at least one clinical treatment.

Our results suggest that personality traits, namely neuroticism and extraversion, are relevant markers of tinnitus distress over time and could be used to statistically distinguish patient groups with clinically relevant changes of tinnitus distress. These markers could inform both treatment responses from clinical studies and future choices on more efficient individual tinnitus treatments.

## Introduction

Tinnitus is a condition characterized by the subjective perception of sounds without an external source^1^. The prevalence of tinnitus is estimated between 10 and 15% for the western societies^2^. Currently, there is no treatment available to reliably and effectively suppress the phantom perception in chronic (i.e., lasting more than six months)^2^, idiopathic presentation of tinnitus. In those cases, treatment focus on the condition’s management, but response to treatments vary considerably across patients. It is yet not fully understood why clinically available treatments do not reliably and effectively suppress the distress associated with tinnitus, but heterogeneity may be a relevant explaining factor^3^. Previous studies suggested that treatments’ low evidence levels could be explained by individual factors^3–5^, and whether personality could be one of such factors is not yet known.

The construct of personality can be described as the individual profile in characteristic patterns of thinking, feeling and behaving^6^. Personality research has implications in a wide range of topics, and several instruments have been developed to characterize and quantify different personality aspects. For instance, the five factor model (FFM, also known as the “Five Factor Model” or “Big Five Personality Traits”) is a widely used, hierarchical model that quantifies five traits of personality, namely Agreeableness, Conscientiousness, Extraversion, Neuroticism, and Openness. Each of the five traits comprises three facets, or subtraits, describing different aspects of each trait^7^. A brief description of the five traits and 15 facets can be found in table 2, and a more in-depth analysis of the BFI2 and its constructs can be found in Soto and John^8^. Apart from the childhood and teenage years, personality is believed to be stable over time, especially after the age of 30^9^. It is also known that the stability of traits decreases with longer retest periods. However, previous studies showed high stability of personality traits over time, with scales presenting r = 0.77 and r = 0.73 in the 6 and 12 years retest interval period^10^.

**Table 1.**
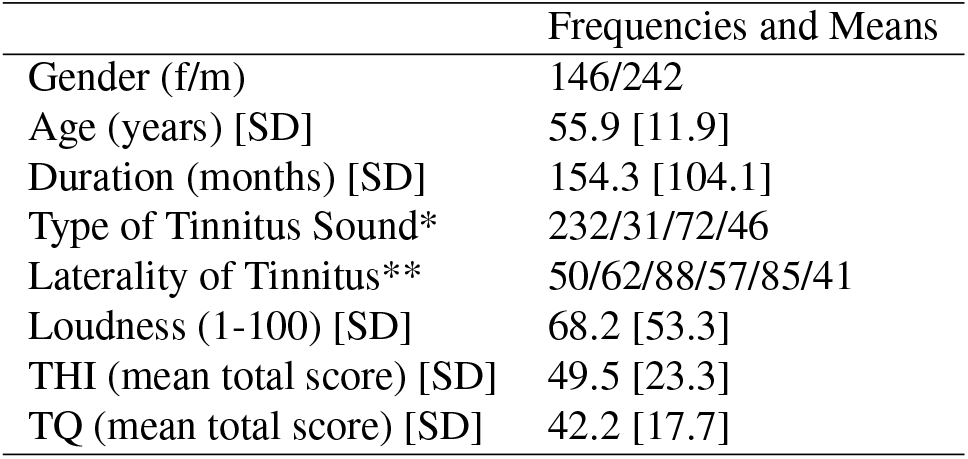
Demographics of our sample. * from left to right: Tonal, Noise, Crickets, Other. ** from left to right: right ear, left ear, both ears (worse in right), both ears (worse in left), both ears (equally bad), inside the head

**Table 2.** Example items of facets in the BFI-2. (R) indicates reversed-keyed items.

| Traits and Facets | Example items |
| --- | --- |
| **Neuroticism** |  |
| Anxiety | 19) Can be tense; Rarely feels anxious or afraid (R) |
| Depression | 9) Stays optimistic after experiencing a setback (R); 39) Often feels sad |
| Emotional Volatility | 44) Keeps their emotions under control (R); Is temperamental, gets emotional easily |
| **Extraversion** |  |
| Sociability | 1) Is outgoing, sociable; 46) Is talkative |
| Assertiveness | 6) Has an assertive personality; 51) Prefers to have others take charge (R) |
| Energy level | 11) Rarely feels excited or eager (R); 41) Is full of energy |
| **Agreeableness** |  |
| Compassion | 2) Is compassionate, has a soft heart; 17) Feels little sympathy for others (R) |
| Trust | 37) Is sometimes rude to others (R); 52) Is polite, courteous to others |
| Respectfulness | 12) Tends to find fault with others (R); 27) Has a forgiving nature |
| **Openness** |  |
| Aesthetic Sensitivity | 35) Values art and beauty; 50) Thinks poetry and plays are boring (R) |
| Intellectual Curiosity | 25) Avoids intellectual, philosophical discussions (R); 40) Is complex, a deep thinker |
| Creative imagination | 15) is inventive, finds clever ways to do things; 30) Has little creativity (R) |
| **Conscientiousness** |  |
| Organization | 3) Tends to be disorganized (R); 33) Keeps things neat and tidy |
| Productiveness | 8) Tends to be lazy (R); 38) Is efficient, gets things done |
| Responsibility | 13) Is dependable, steady; 28) |

Looking at tinnitus specifically, Langguth and colleagues^11^ used the FFM to describe the personality traits of tinnitus patients. The authors found that tinnitus patients tend to score higher in neuroticism, and lower in agreeableness. Additionally, a recent scoping review^12^ suggested that personality traits, such as high neuroticism and low extraversion, are common hallmarks of tinnitus patients. Previous works have investigated the prevalence of of the “type D” personality and tinnitus. Type D personality is characterized by both social inhibition and neuroticisms, and has been shown to be prevalent in tinnitus patients^13,14^.

However, studies assessing the putative role of tinnitus distress longitudinally are scarce. Recently, Kleinstäuber and colleagues^15^ investigated the role of personality traits on internet-delivered cognitive behavior therapy (iCBT) in chronic tinnitus patients. The results indicated that different traits can predict the outcome of an iCBT intervention after different time periods (e.g., 3, 6, 12 months after treatment), underscoring the often overlooked influence of personality on treatment outcomes in tinnitus. However, some questions remain unanswered: It is yet not clear if the effects of personality can predict tinnitus-related distress over time, disregard of whether a patient tried any type of treatment or not. It is also unclear whether personality mediates the outcome of psychological-based interventions or, in general, mediates all kinds of tinnitus-related interventions. From a clinical perspective, these open questions are of utmost importance to better understand differences in clinically relevant changes of tinnitus symptomatology. In the study at hand, we aimed at 1) replicating the previous results obtained by Langguth and colleagues^11^, but with a larger sample size; 2) investigating which facets of relevant personality traits account for tinnitus distress; 3) investigating the role of personality traits on tinnitus distress over time; and, of central interest, 4) evaluating whether such traits may be of clinically relevance to the treatment response.

Our hypotheses were: 1) neuroticism correlates positively with tinnitus distress over time (i.e., the higher neuroticism is, the lower distress tends to decline over time), whereas 2) extraversion correlates negatively with tinnitus distress over time; 3) neuroticism correlates positively with changes in distress over time, and 4) neuroticism and extraversion inform differences in clinically relevant grading of the tinnitus distress questionnaires.

## Results

### Cross-sectional analysis of personality traits and facets

Demographics and tinnitus characteristics of our sample can be found in table 1. First, we applied multiple linear regressions with personality traits as independent variables and the scores of THI and TQ at T1 as dependent variables. Table 3 shows both statistical models. Both models showed a negative association between extraversion and the questionnaire outcomes, and a positive association between neuroticism and the questionnaires outcomes. Moreover, the models could explain 25% of the variance (R2 = 0.25, F(5,331) = 23.47, p < 0.001) with the THI as the dependent variable and 24% of the variance (R2 = 0.24, F(5,331) = 22.69, p < 114 0.001) with the TQ as the dependent variable.

**Table 3.** Linear regression models with personality traits as independent variables and THI and TQ at T1 as dependent variables

|  | TQ |  |  |  | THI |  |  |  |
| --- | --- | --- | --- | --- | --- | --- | --- | --- |
|  | coefficients | Std. Error | t value | p value | coefficients | Std. Error | t value | p value |
| Intercept | 19.33 | 10.99 | 1.76 | 0.08 | 4.44 | 14.17 | 0.31 | 0.75 |
| Agreeableness | 0.17 | 0.16 | 1.08 | 0.28 | 0.32 | 0.20 | 1.57 | 0.12 |
| Conscientiousness | 0.09 | 0.13 | 0.71 | 0.48 | -0.01 | 0.17 | -0.05 | 0.96 |
| Extraversion | -0.34 | 0.13 | -2.64 | **0.01** | -0.35 | 0.17 | -2.09 | **0.04** |
| Neuroticism | 0.87 | 0.12 | 7.53 | **<0.01** | 1.27 | 0.15 | 8.53 | **<0.01** |
| Openness | -0.19 | 0.12 | -1.56 | 0.12 | -0.03 | 0.16 | -0.21 | 0.84 |

We modeled the facets of extraversion and neuroticism as independent variables and the scores from T1 as the dependent variable as these two traits were statistically significant in the previous models. Results are presented in tables 4 and 5. Regarding the facets of neuroticism, both the models showed a statistical significant association between depression (positive) and emotional volatility (negative) with the dependent variable. Anxiety was statistically significantly associated with the TQ scores. The effect size of of both models with THI and TQ as dependent variables was 15% (R2=.15, F(3, 333) = 20.78, p < 0.001) and 16% (R2=.16, F(3, 335) = 22.87, p < 0.001) respectively. Regarding the facets of extraversion, the model with the THI as a dependent variable showed a negative significant statistical association with the facet “energy level”. The models with THI and TQ as dependent variables and extraversion facets as independent variables could explain, respectively, 2% (R2=.02, F(3,333) = 2.78, p < 0.05), and 1% of the variance (R2=.01, F(3,335) = 2.37, p = 0.07).

**Table 4.** Linear regression models with the facets of the personality trait “neuroticism” as independent variable and THI and TQ at T1

|  | TQ |  |  |  | THI |  |  |  |
| --- | --- | --- | --- | --- | --- | --- | --- | --- |
|  | coefficients | Std. Error | t value | p value | coefficients | Std. Error | t value | p value |
| Intercept | 35.09 | 5.60 | 6.27 | 0.00 | 37.75 | 7.30 | 5.17 | 0.00 |
| Anxiety | -1.37 | 0.64 | -2.13 | **0.03** | -1.35 | 0.84 | -1.60 | 0.11 |
| Depression | 3.45 | 0.48 | 7.22 | **<0.01** | 4.19 | 0.62 | 6.73 | **<0.01** |
| Emotional Volatility | -1.40 | 0.47 | -2.95 | **<0.01** | -1.75 | 0.62 | -2.84 | **<0.01** |

**Table 5.**
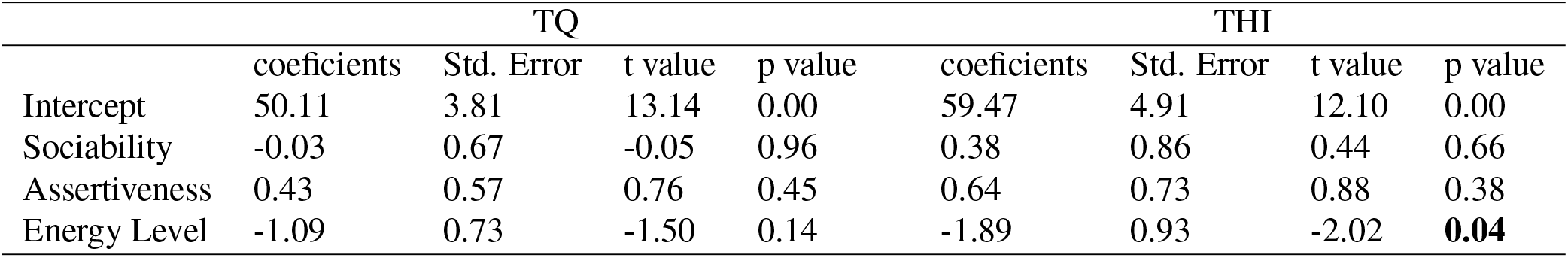
Linear regression models with the facets of the personality trait “extraversion” as independent variable and THI and TQ at T1

|  | TQ |  |  |  | THI |  |  |  |
| --- | --- | --- | --- | --- | --- | --- | --- | --- |
|  | coefficients | Std. Error | t value | p value | coefficients | Std. Error | t value | p value |
| Intercept | 50.11 | 3.81 | 13.14 | 0.00 | 59.47 | 4.91 | 12.10 | 0.00 |
| Sociability | -0.03 | 0.67 | -0.05 | 0.96 | 0.38 | 0.86 | 0.44 | 0.66 |
| Assertiveness | 0.43 | 0.57 | 0.76 | 0.45 | 0.64 | 0.73 | 0.88 | 0.38 |
| Energy Level | -1.09 | 0.73 | -1.50 | 0.14 | -1.89 | 0.93 | -2.02 | **0.04** |

### 1 Difference in tinnitus distress Between T1 and T2

Next, we modelled a multiple linear regression with personality traits as independent variables and the difference in the THI and TQ between T2 and T1 as dependent variables. Since T1 represents a visit between 2012 and 2017, we added year of visit as an independent variable in the models. This way, we could account for a potential cumulative effect of time over tinnitus distress over time. The results are presented in table 6. No personality trait reached statistical significance in either models. Yet, we can report statistical trends for the neuroticism in the TQ (t = 1.68, p = 0.09), and for conscientiousness in the THI (t = −1.8, p = 0.07. The model with the THI as dependent variable had 5% of the variance explained by the predictors (R2 = .05, F(6,327) = 3.647, p < 0.01), whereas the model with the TQ as the dependent variable had 3% of the variance explained (R2 = .03, F(6,331) = 2.569, p = 0.01).

**Table 6.** Linear regression models with the personality traits as independent variable and difference between the scores of THI and TQ in T2 - T1

|  | TQ |  |  |  | THI |  |  |  |
| --- | --- | --- | --- | --- | --- | --- | --- | --- |
|  | coefficients | Std. Error | t value | p value | coefficients | Std. Error | t value | p value |
| Intercept | -1863.00 | 737.30 | -2.53 | 0.01 | -2470.00 | 1027.00 | -2.41 | 0.02 |
| agreeableness | 0.07 | 0.13 | 0.54 | 0.59 | 0.09 | 0.18 | 0.51 | 0.61 |
| conscientiousness | -0.16 | 0.10 | -1.57 | 0.12 | -0.26 | 0.14 | -1.80 | 0.07 |
| extraversion | -0.03 | 0.10 | -0.26 | 0.79 | -0.19 | 0.14 | -1.36 | 0.17 |
| neuroticism | 0.16 | 0.09 | 1.68 | 0.09 | 0.18 | 0.13 | 1.37 | 0.17 |
| openness | 0.00 | 0.10 | 0.01 | 0.99 | -0.03 | 0.14 | -0.25 | 0.80 |
| year of visit | 0.92 | 0.37 | 2.52 | **0.01** | 1.23 | 0.51 | 2.41 | **0.02** |

**Table 7.** Linear regression models with personality traits as independent variables and THI and TQ at T1 as dependent variables

|  | TQ |  |  |  | THI |  |  |  |
| --- | --- | --- | --- | --- | --- | --- | --- | --- |
|  | coefficients | Std. Error | t value | p value | coefficients | Std. Error | t value | p value |
| Intercept | 11.5 | 11.4 | 1.03 | 0.3 | 2.29 | 13.56 | 0.169 | 0.86 |
| Agreeableness | 0.24 | 0.16 | 1.48 | 0.13 | 0.45 | 0.20 | 2.3 | 0.02 |
| Conscientiousness | -0.07 | 0.13 | -0.55 | 0.58 | -0.23 | 0.16 | -1.45 | 0.13 |
| Extraversion | -0.34 | 0.13 | -2.59 | 0.01 | -0.51 | 0.16 | -3.26 | <0.01 |
| Neuroticism | 1.06 | 0.12 | 8.77 | <0.001 | 1.49 | 0.15 | 10.45 | <0.001 |
| Openness | -0.19 | 0.12 | -1.51 | 0.13 | -0.17 | 0.15 | -0.7 | 0.48 |
| F Statistic | F(5, 338) = 30.63 |  |  |  | F(5, 336) = 42.12 |  |  |  |
| Adjusted R squared | 0.31 |  |  |  | 0.38 |  |  |  |

### 2 Clinical significant differences

To further explore the potential role of personality traits on tinnitus distress over time, we grouped patients into three groups based on the difference between the scores of the THI and TQ on T2 - T1. The results are presented in figures 1 and 2. Neuroticism was statistically significantly lower in patients in the groups “improved” and “stable” compared to the “worsened” group in the THI (t = −3.3, p-value < 0.01, d = 0.5; t = −2.06, p-value = 0.04, d = 0.34). Likewise, patients in the “stable” groups in the TQ showed statistically significant lower neuroticism scores than the patients binned in the “worsened” group (t = −2.26, p-value = 0.03, d = 0.45). Whereas higher neuroticism was associated with worsening in tinnitus, higher extraversion was associated with improvement in tinnitus distress. Regarding the THI, the patients binned in the “improvement” group scored higher than patients binned in the “stable group” (W = 2.55, p-value = 0.01, d = 0.31) and higher than patients binned in the “worsened” group (t = 2.2, p-value = 0.03, d = 0.34).

**Figure 1.**
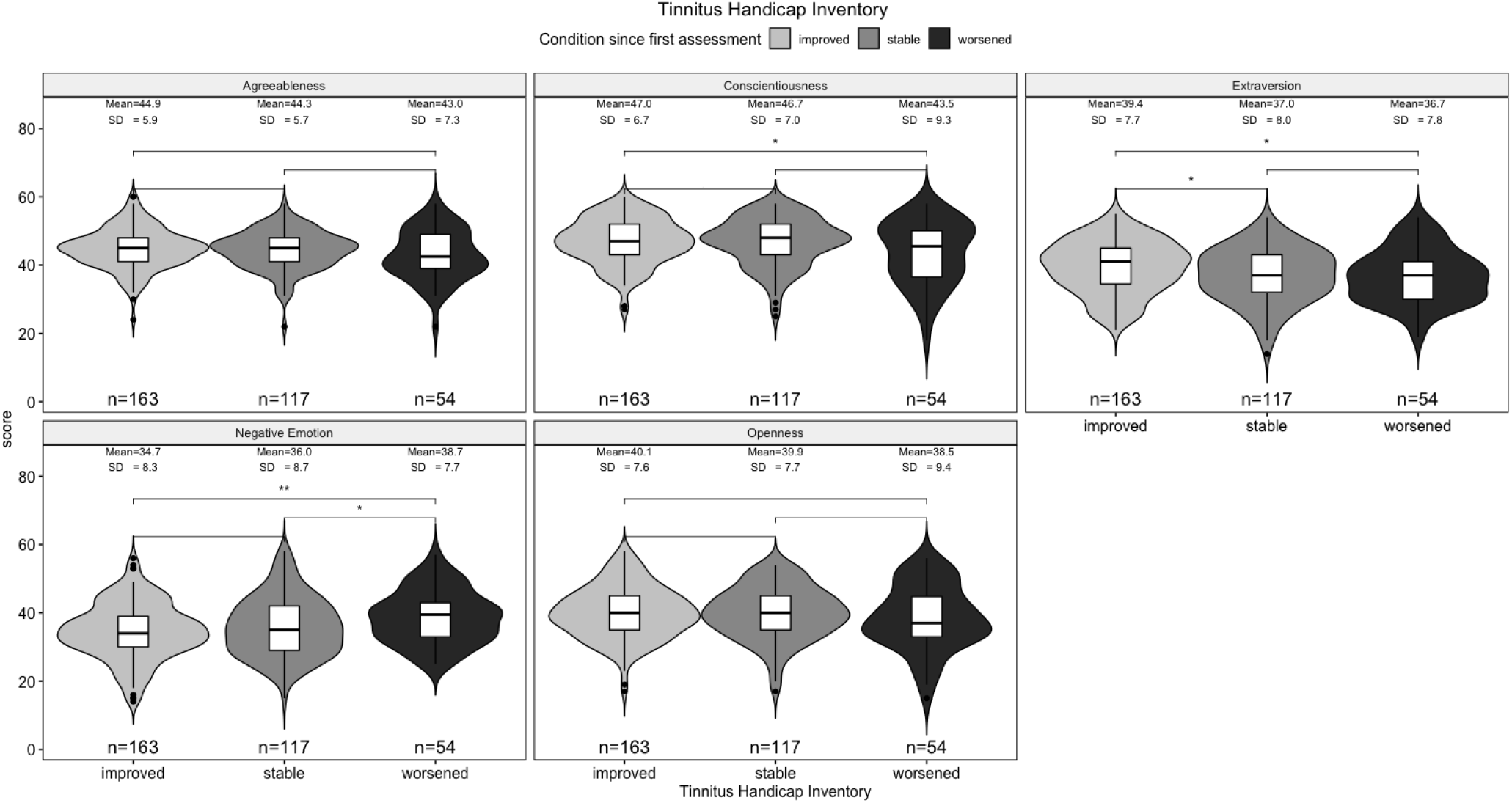
THI change grades and personality traits.

**Figure 2.**
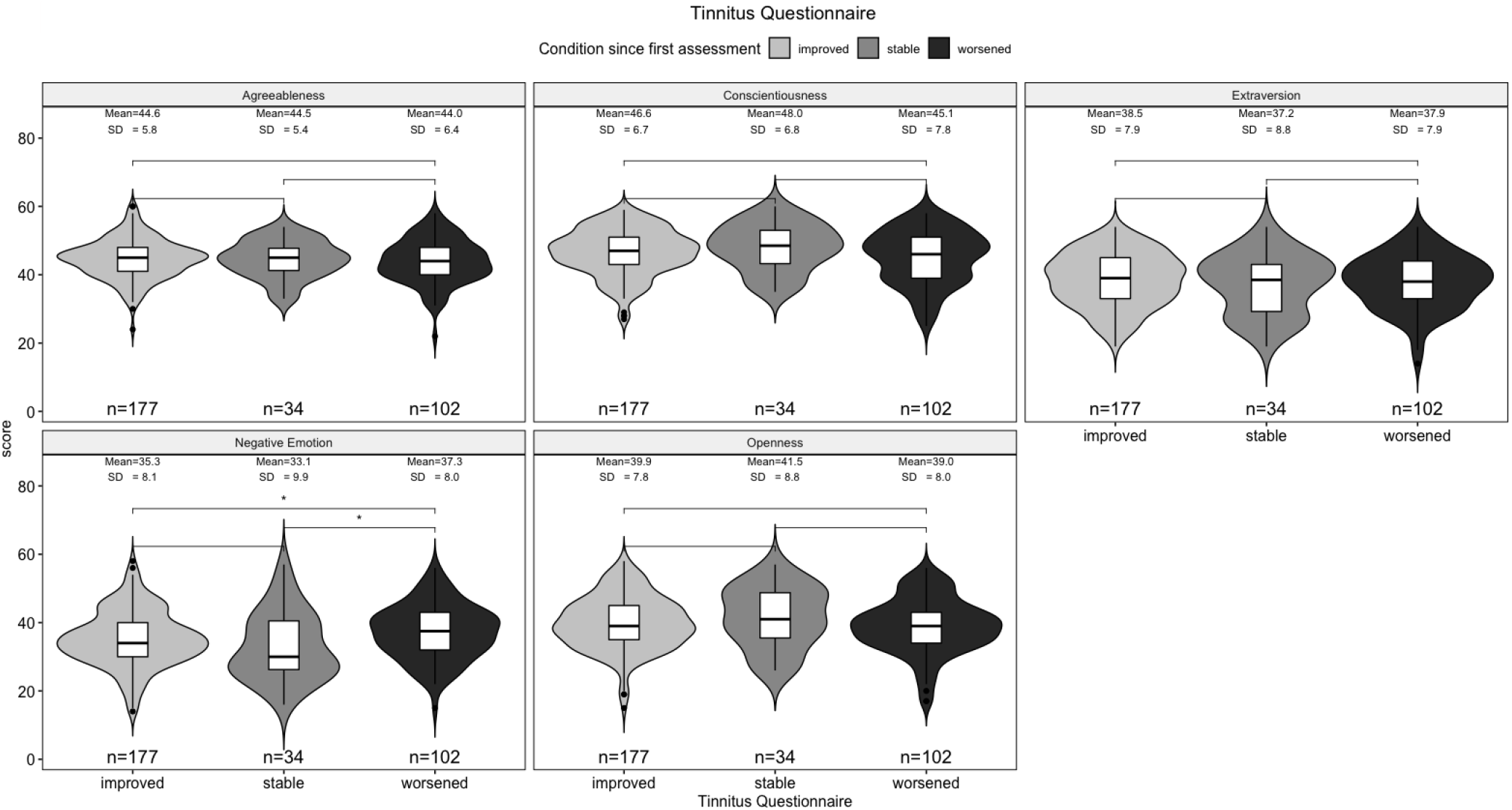
TQ change grades and personality traits.

### 3 Clinically relevant differences and treatments

For this analysis, we clustered patients in two groups: patients who tried at least one tinnitus-related treatment between T1 and T2, and patients who did not. Results are presented in figures 3 and 4. Patients who did not try any treatment between T1 and T2, and binned in the “improved” group for the THI scored lower in neuroticism than patients binned in the “worsened” group (W = 2737.5, p-value = 0.01165). Similar results were observed with the TQ (W = 6105, p = 0.02), with the addition of a statistical significant difference between the groups “stable” and “worsened” (W = 159 895.5, p = 0.03). Regarding extraversion, we observed statistical significant differences between the “improved” and “stable” groups (W = 8372.5, p-value < 0.01, d = 0.37), and between the “improved” and “worsened” group (4418.5, p-value = 0.02, d = 0.39) for the THI in the group of patients who did not try any treatment between T1 and T2. For the TQ, we observed a statistical significant change between the groups “improved” and “worsened” among patients that tried at least one treatment between T1 and T2 (W = 51, p-value = 0.03).

**Figure 3.**
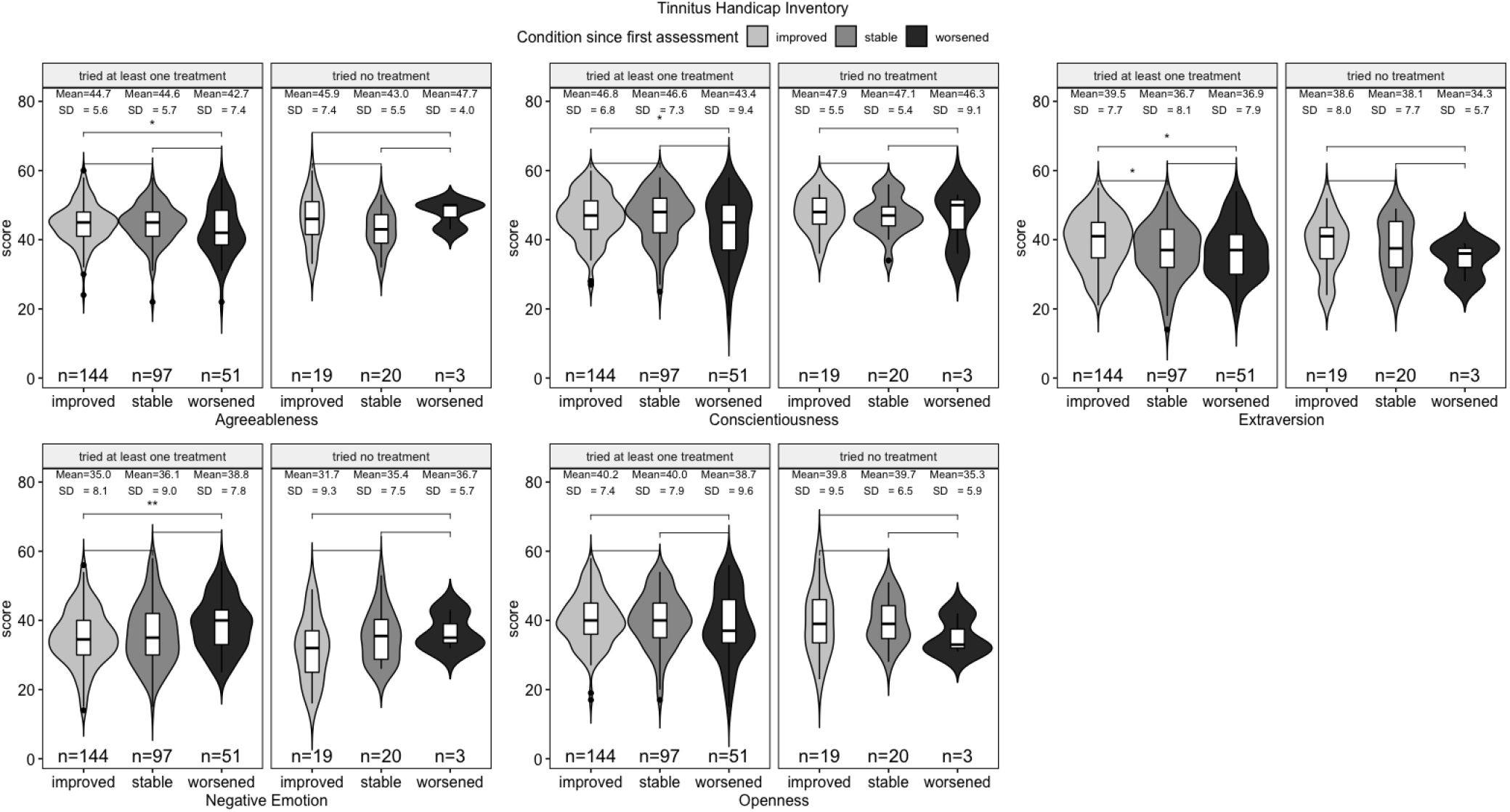
THI change grades and personality traits.

**Figure 4.**
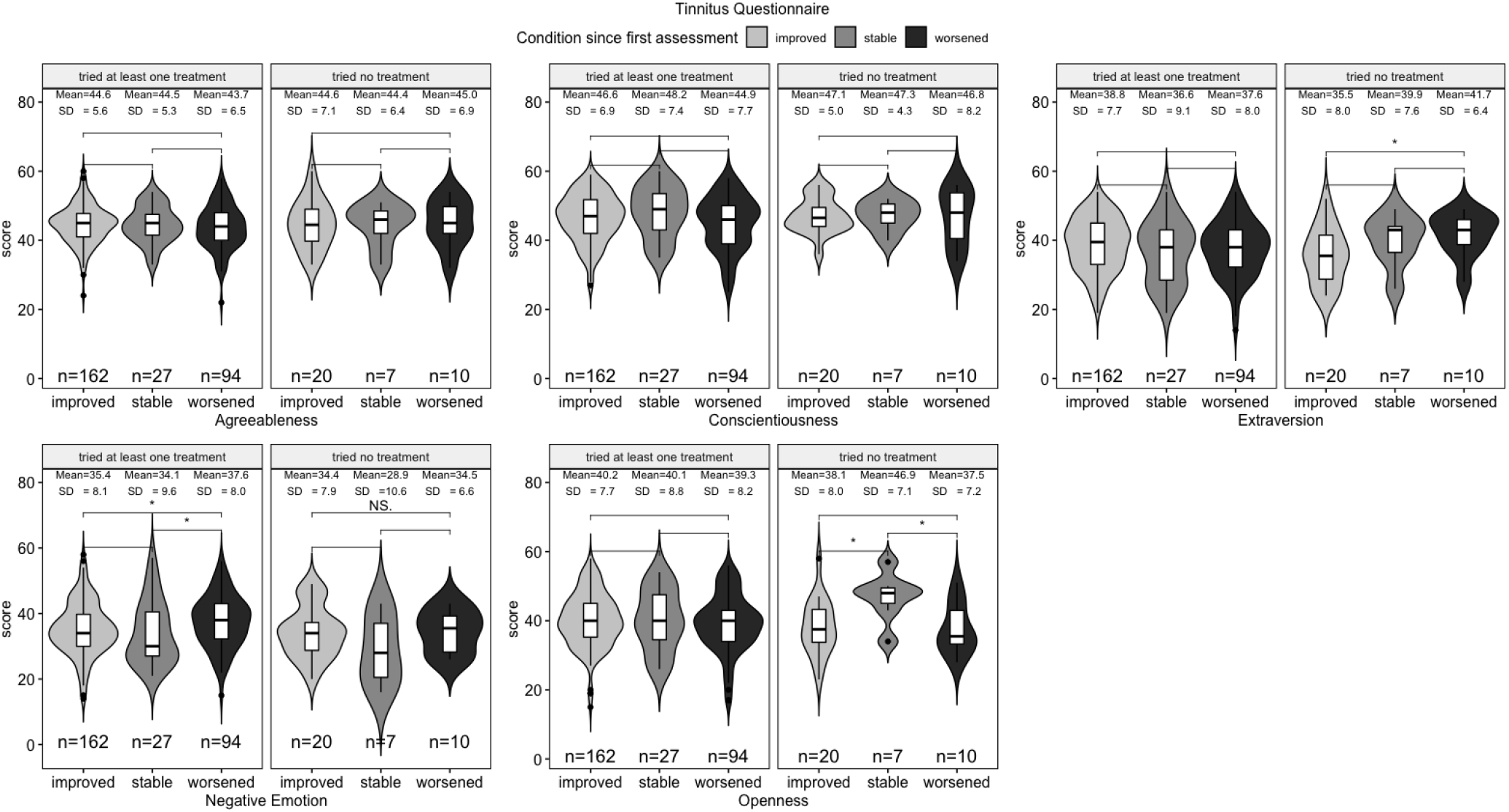
TQ change grades and personality traits.

## Discussion

The present study was the first to systematically analyze the effect of personality on the longitudinal trajectory of tinnitus distress. We showed that neuroticism is related to changes in tinnitus distress, i.e., clinically relevant changes in the grade of tinnitus distress as measured by TQ and THI. More specifically and of central interest, neuroticism was higher in patients with worsened clinical status compared to patients with improved clinical status (measured with THI as well as TQ). Looking at extraversion, the group showing clinical improvement exhibited significantly higher scores compared to the groups of stable and worsened clinical status (measured with THI but not significant for TQ).

Our cross-sectional analysis between personality and tinnitus distress partially reproduced the findings of^11^, as neuroticism showed a positive association with tinnitus distress. However, we could only observe an association between agreeableness and the THI. Our results are in line with a scoping review looking systematically at personality traits and tinnitus distress, as lower extraversion and higher neuroticism scores were associated with increased THI and TQ^12,16^.

To the best of our knowledge, our study is the first one to investigate facets of the FFM in relation to tinnitus distress. We identified two facets of neuroticism, namely depression and emotional volatility, as significant predictors of the THI and TQ at T1. We also identified energy level, a facet of extraversion, to be a a significant predictor of the THI. Interestingly, those facets are characteristic of the “type D” personality, which is prevalent among high distressed tinnitus patients^12,14^. These results may open new venues of investigation, especially to better understand the relation between depression and tinnitus. A previous review^16^ discussed the challenges of separating depression and tinnitus, as the symptoms presented by both groups and the underlying mechanisms between the two conditions are often convoluted (e.g., neuronal mechanisms^17^). Future studies could investigate the relation between the two conditions by comparing acute depression (e.g., as measured by the Beck’s or Major Depression Inventory), depressive personality (e.g., as measured by the “depression” facet) and biomakers of depression, such as the brain-derived neurotrophic factor^18^.

We could not observe a significant effect of any of the five personality traits on tinnitus distress over time in our multiple linear regression setup (except a trend for neuroticism and TQ, p<0.09). On the other hand, the year of first assessment was a significant predictor of the difference in the THI and TQ in T2 - T1, suggesting that the longer the period between T1 and T2, the greater the decrease in tinnitus distress. To test for clinical relevance, which surpasses mere psychometric statistical analysis and therefore generate important practical insights for clinical routine in tinnitus, we grouped patients in three groups based on previous literature^19,20^: “clinically improved”, “clinically stable”, and “clinically worsened”.

These differences reflected the difference in the THI and TQ between two time points. Patients grouped in the “worsened” group had higher levels of neuroticism than the other two groups in both the THI and the TQ, and extraversion was significantly higher in the “improved” group than in the other two for the differences in the THI but not in the TQ. Both findings are aligned with previous cross-sectional literature, as low extraversion and high neuroticism seem to be associated with higher tinnitus distress^12^. Additionally, these differences in personality related to clinically relevant changes in tinnitus distress may have prognostic value, as they were able to statistically distinguish three clinically relevant groups of tinnitus patients. Similar effects were observed when we divided the three groups between patients who tried at least one clinical trial for their tinnitus and those who did not. Our results suggests that personality plays a role on the change of distress among patients who tried at least one treatment. There is an on going debate about the role of personality on placebo effects in clinical trials, and researchers are increasingly aware of its putative effects alongside other factors such as positive/negative expectations and patient-clinician relationship^21^. We recommend future tinnitus studies to measure and report personality traits in their clinical trials, as personality could either represent a confounder in trials’ outcomes or be used to identify efficient individual treatments.

Tinnitus heterogeneity has been implicated as a major obstacle to improving the condition’s management, and thus tinnitus subtyping has been settled as a major objective in the research community^4,5^. To the best of our knowledge, no study systematically investigated the putative effects of personality on treatment outcomes in tinnitus, and it is yet unknown whether personality is a component of tinnitus heterogeneity. Previous studies have shown that personality mediates the outcomes of internet-delivered Cognitive Behavior Therapy^15^ and Lidocaine treatment^22^.

Our results support these findings, and future research should further explore the role of personality on tinnitus heterogeneity and its implications on treatment outcomes. For instance, although the relation between personality and coping strategies is well characterized in the overall literature^23^, little is known about it in tinnitus patients. Coping strategies vary per individual, and tend to be influenced by personality^23^. We recommend future research to explore the relation between personality, coping, and its potential effects on neuroticism is often associated with avoidance behavior, which, in turn, is implicated in higher anxiety and distress in tinnitus patients^24^.

This study has inherent limitations. First, we collected the FFM at T2 and not at T1. Whether personality is a crystallized construct, ie., it does not change over time, is debatable. For instance, it is possible to change FFM scores through experimental manipulation^25^. Conversely, Cost and Mccrae^9,10^ reviewed the evidence for the stability of the construct longer periods of time. The stability of personality over time, in our cohort up to seven years to patients who visited our clinic in 2012, is an important assumption of our analysis here^10^. As proposed above, it would therefore be helpful to assess personality within standard psychometric tinnitus batteries at all time points in clinical trials or general longitudinal research. Second, we could not properly investigate the role of personality among patients who did not try any treatment due to small sample size. Whether clinically significant tinnitus habituation can be explained by personality remains a relevant, open question. Third, we could not discard a potential bias among those who responded to our survey at T2. To the best of our knowledge, no former study investigated whether patients participate in surveys and/or clinical trials. However, apart from a 2-point difference in the TQ, we could not find statistically significant differences between the sample which responded the survey and the sample which did not.

In conclusion, our results suggest that personality traits, namely neuroticism and extraversion, can explain a large portion of the variance of tinnitus distress. Those two traits are relevant markers of tinnitus distress over time and can be used to statistically distinguish patient groups with clinically relevant changes of tinnitus distress. Personality assessments could provide valuable information to clinicians and researchers, and may eventually be used to deliver personalized treatments for tinnitus patients. Future studies would furthermore profit from assessing personality at several time points to further investigate interactions between personality and tinnitus.

## Methods

### 3.1 Participants

Previous patients of the Tinnitus Outpatient Center of the University of Regensburg were invited to participate in this questionnaire survey by a letter with questionnaires and consent forms. 1213 letters were sent to patients who visited the clinic between 2012 and 2017, from which 388 sent back with signed consent forms. The study was approved by the ethical committee at the faculty of medicine of the University of Regensburg (study number 18-1041-101).

### 3.2 Data collection

The longitudinal analysis considered the first questionnaire assessment during the visit to the clinic as T1 and the assessment with the questionnaires delivered by mail in 2018 as T2. The THI, TQ and TSCHQ were collected at T1 and T2, and the BFI-2 and an informal in-house questionnaire asking patients which treatments they tried between T1 and T2. Regarding the BFI-2, patients filled in a previously validated German version of the questionnaire (Danner et al., 2016).

### 3.3 Statistical analysis

To evaluate the potential predictive role of the BFI2 on tinnitus distress over time, we grouped patients in three categories: clinically “improved”, “stable”, and “worsened”. The groups reflect the difference of the THI and TQ scores as the difference in scores between the two time points, and followed the guidelines of Zeman et al. (2011) and Adamchic et al. (2012). More specifically, for the THI, a decrease in >7 points at T2 was considered a significant clinical improvement, an increase in >7 was considered a clinical significant worsening, and the patients who did not score at T2 higher or lower than seven points were grouped as “stable”. For the TQ, the thresholds for improvement and worsening were, respectively, 5 points lower and 1 point higher in T2 compared to T1. Since T1 spanned between 2012 and 2017, the year of visit T1 was treated as covariate in the longitudinal analysis. All statistical analyses were conducted with R statistical software (version 3.4.4, R Development Core Team, 2008), alongside the “tidyverse” package (Wickham, 2017). Effect sizes were calculated with the package “effsize” (Torchiano, 2018), and plots were generated with the “ggpubr” package. Non-parametric tests were used when test assumptions were not met. P values below 0.05 were considered statistically significant.

## Data Availability

The data is not publicly available.
Enquiries should be directed to the corresponding author.

## Author contributions statement

PN, DF, WS, and JS defined the study design and interpreted the results.

BL, PN, MS, and WS interpreted the results, and provided critical feedback during the manuscript preparation.

DF, MS and JS were responsible for data collection.

## Additional information

The authors declare no conflict of interest.

The corresponding author is responsible for submitting a competing interests statement on behalf of all authors of the paper.

This statement must be included in the submitted article file.

